# The Relationship Between Symptoms and Job Loss among Japanese Workers During the COVID-19 Pandemic: A Prospective Cohort Study

**DOI:** 10.1101/2022.09.06.22279656

**Authors:** Shintaro Okahara, Yoshihisa Fujino, Tomohisa Nagata, Mami Kuwamura, Kosuke Mafune, Keiji Muramatsu, Seiichiro Tateishi, Akira Ogami, Koji Mori, the CORoNaWork project

## Abstract

**Objectives:** The aim of this study was to clarify which workers’ symptoms led to unemployment during the COVID-19 pandemic.

**Methods:** This was a prospective cohort study using questionnaires about COVID-19 administered to Japanese workers. A baseline survey conducted in December 2020 was used to determine workers’ health history. Unemployment since the baseline survey was ascertained with a follow-up survey in December 2021. The odds ratios (ORs) of unemployment were estimated using a multilevel logistic model with adjusted covariates nested in prefecture of residence.

**Results:** Males (*n* = 8,682) accounted for 58.2% of the total sample (*n* = 14,910), and the mean age was 48.2 years. Multivariate analysis showed that workers with “pain-related problems,” “limited physical movement and mobility,” “fatigue, loss of strength or appetite, fever, dizziness, and moodiness,” “mental health problems,” or “sleep” had a greater probability of resigning for health reasons, resigning for all reasons other than retirement, and being unemployed. Those with “skin, hair, and cosmetic concerns” or “eye-related matters” had a greater probability of becoming unemployed.

**Conclusions:** We identified an association between workers’ symptoms and resignation or unemployment, with different symptoms having different ORs. Furthermore, there were differences in the associations among the effects of symptoms, work dysfunction, resignation/unemployment, and attitudes of others. Preventive interventions to help workers resolve or improve their symptoms could prevent their becoming unemployed or resigning.

## Introduction

It is important to provide opportunities for workers with health conditions or illnesses to continue to work, not only to enhance their quality of life, but also to increase labor productivity and reduce the social security burden. Japan is facing a significant problem in the form of a declining labor force as a result of the falling birthrate and aging population. Thus, it is necessary to induce groups that currently cannot work to enter the labor force. For example, females, older people, and those unable to work for health reasons should be encouraged to participate in the labor market. It is also necessary to improve the labor productivity of workers currently participating in the labor market. The Japanese government introduced a policy of “harmonizing work with disease treatment and prevention” with the aim of creating a working and social environment in which workers with health problems can work stably over the long term^1^.

Workers with health problems face various difficulties in the workplace. A worker with a health problem might require short-term, long-term, or repeated absences from work as a result of either the need for medical treatment or their symptoms^2-5^. They face both quantitative and qualitative obstacles in relation to their work performance as a result of their symptoms and the side effects of treatment^2,5-7^, and can experience stigma related to their health problems in the workplace^8,9^. Thus, workers with health problems are likely to experience unemployment^10,11^ and remain unemployed for long periods of time^12^.

The COVID-19 pandemic has affected workers’ disease management. Initially, it led to interruptions in treatment for those with chronic illnesses^13^ because of concerns about COVID-19 infection, scheduling changes, restrictions on outpatient access as a result of limited medical resources, and people’s economic instability. The pandemic also affected people’s lifestyles with the introduction of lockdowns, social distancing, and working from home in an attempt to curb the spread of COVID-19, resulting in people becoming less physically active^14,15^. As people’s levels of interaction decreased, their levels of stress, loneliness, and depression increased^16-18^, and alcohol consumption and smoking increased^19-21^.

The COVID-19 pandemic has created precarious employment conditions. In Japan, in an effort to prevent the spread of COVID-19, the economic activities of companies have been severely restricted, and people’s consumption has also declined. This has had a significant impact on the Japanese economy, as evidenced by negative GDP growth^22^. In terms of employment and labor conditions, there was a significant drop in the number of employees, as well as a decrease in working hours and wages^23^. During the COVID-19 pandemic, about 11% of Japanese workers who required regular treatment experienced treatment interruption. Disadvantageous socioeconomic status, poor health, and unfavorable lifestyle habits were associated with treatment interruption^13^.

Even prior to the COVID-19 pandemic, workers with health problems were more likely to experience unemployment, and this was most likely exacerbated by the COVID-19 pandemic. However, the specific impact of the COVID-19 pandemic is unknown, and thus the aim of this study was to identify the symptoms among workers that led to unemployment during the COVID-19 pandemic.

## Materials and methods

This study was conducted under the Collaborative Online Research on the Novel-coronavirus and Work (CORoNaWork) Project. The details of the study protocol are provided elsewhere^24^. Briefly, we administered a baseline questionnaire in December 2020 and a follow-up questionnaire in December 2021.

For the baseline survey in December 2020, a total of 33,087 workers were recruited throughout Japan from 605,381 randomly selected panelists who were registered with an Internet survey company. The inclusion criteria were currently working and aged 20–65 years, and we did not invite healthcare professionals or caregivers to participate. We used cluster sampling with stratification by sex, job type, and region. We excluded 6,051 invalid responses by the criteria used to determine unreliable responses included extremely short response times (less than 6 minutes), extremely low weight (less than 30 kg), extremely low height (less than 140 cm), inconsistent responses to similar questions throughout the survey (e.g., questions about marital status and region of residence), and questions used to identify fraudulent responses. We distributed the follow-up questionnaire in December 2021 to the 27,036 people with valid responses to the baseline questionnaire, of whom 18,560 responded (a follow-up response rate of 68.6%). We excluded self-employed workers (n=1,635), workers in small/home offices (n=284), agriculture, forestry, and fishery workers (n=146), professionals such as lawyers, tax accountants, and medical practitioners (n=1,137), and workers whose labor contract differed markedly from that of standard workers (n=448). This left a final sample of 14,910 workers.

The study was approved by the Ethics Committee of the University of Occupational and Environmental Health, Japan (reference nos. R2-079 and R3-006). Informed consent was obtained from all participants.

### Explanatory variables

At baseline, we identified the workers’ health conditions by asking them about symptoms known to be strongly associated with presenteeism in previous studies: “Which of the following is closest to the health problem that is most affecting your work?” They answered by selecting one of the following options: “No particular problems”; “Pain”; “Physical movement and mobility”; “Fatigue, loss of strength or appetite, fever, dizziness, and moodiness”; “Toileting and defecation”; “Mental health”; “Skin, hair, and cosmetic concerns”; “Sleep”; “Eye-related matters”; “Nasal matters”; “Hearing”; or “Other”^25^.

### Outcomes

Resignation and unemployment were ascertained as follows. First, the baseline survey was limited to people who were employed at the time. In the follow-up survey, in response to the question “Have you retired (or changed jobs) since December 2020?” respondents were asked to select one of the following six options: “I have not retired (or changed jobs) at all”; “I have retired (or changed jobs) for health reasons”; “I have retired (or changed jobs) because of company downsizing, termination of employment, or expiration of contract”; “I have retired (or changed jobs) because of bankruptcy, or business closing”; “I have retired because of mandatory retirement”; or “I have retired (or changed jobs) for other reasons.” If the respondent answered that “I have retired (or changed jobs) for health reasons,” this was defined as “resigned for health reasons.”

The follow-up survey also asked whether there had been a period of unemployment since December 2020. If the participants had experienced unemployment between the baseline and follow-up surveys, this was defined as “experienced unemployment.”

### Control variables

We retrieved the following data from the baseline survey for inclusion as control variables: age, sex, marital status, annual household income, education, job type, company size (number of workers), smoking status, and alcohol consumption habits. Age was treated as a continuous variable. Marital status was classified into three categories as follows: married, divorced or widowed, and never married. Annual household income was classified into four categories as follows: less than 4 million Japanese yen (JPY), 4.00–5.99 million JPY, 6.00–7.99 million JPY, and 8 million JPY or more. Education was classified into three categories as follows: junior high school, high school, and vocational school/college, university, or graduate school. Job type was classified into three categories as follows: desk work, work involving communicating with people, and manual work. Company size (number of workers) was classified into four categories as follows: less than 30, 30–99, 100–999, and 1000 or more. Smoking status was categorized into two categories as follows: current smoker or nonsmoker. Alcohol consumption habits were categorized into three categories as follows: consuming alcohol on 4 or more days per week; consuming alcohol on 3 or fewer days per week; and rarely or never consuming alcohol.

### Statistical analyses

In the analyses, health conditions were treated as the exposure variables, and resignation or unemployment were treated as the outcome variable. The odds ratios (ORs) of experiencing resignation or unemployment associated with each health condition were estimated using a multilevel logistic model, which was nested in the prefecture of residence to account for regional differences. Age/sex-adjusted and multivariate-adjusted ORs were estimated. The multivariate model included age, sex, marital status, annual household income, education, job type, company size (number of workers), smoking status, and alcohol consumption habits, and the rate of incidence of COVID-19 by prefecture at baseline. A p value of less than 0.05 was considered statistically significant. All analyses were conducted using Stata (Stata Statistical Software: Release 17; StataCorp LLC, TX, USA).

## Results

The baseline characteristics of the respondents are shown in Table 1. The sample of 14,910 included 8,682 males (58.2%) and the mean age was 48.2 years. A total of 475 participants (3.2%) resigned for health reasons, and 2430 participants (16.3%) experienced unemployment.

**Table 1.**
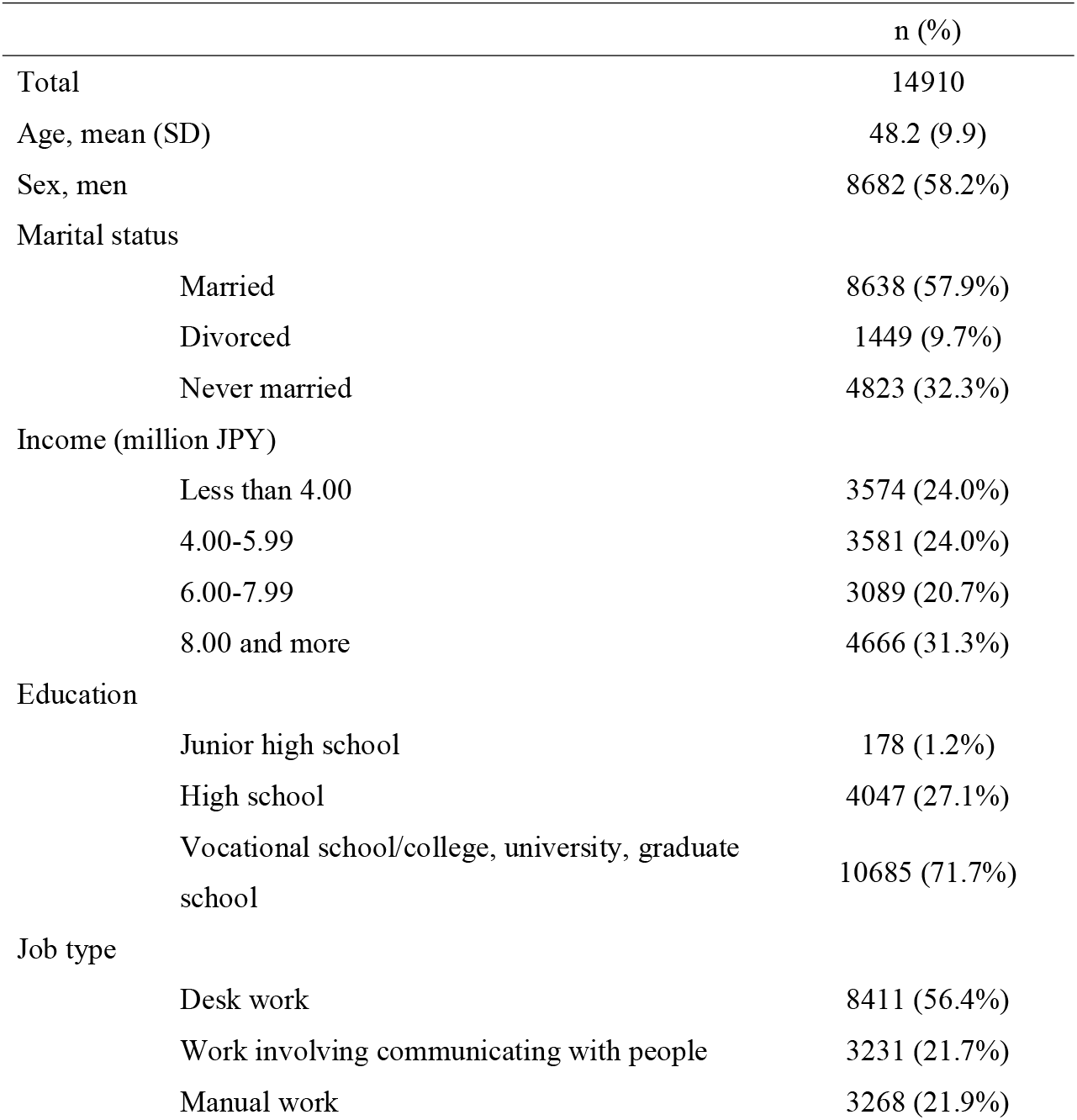
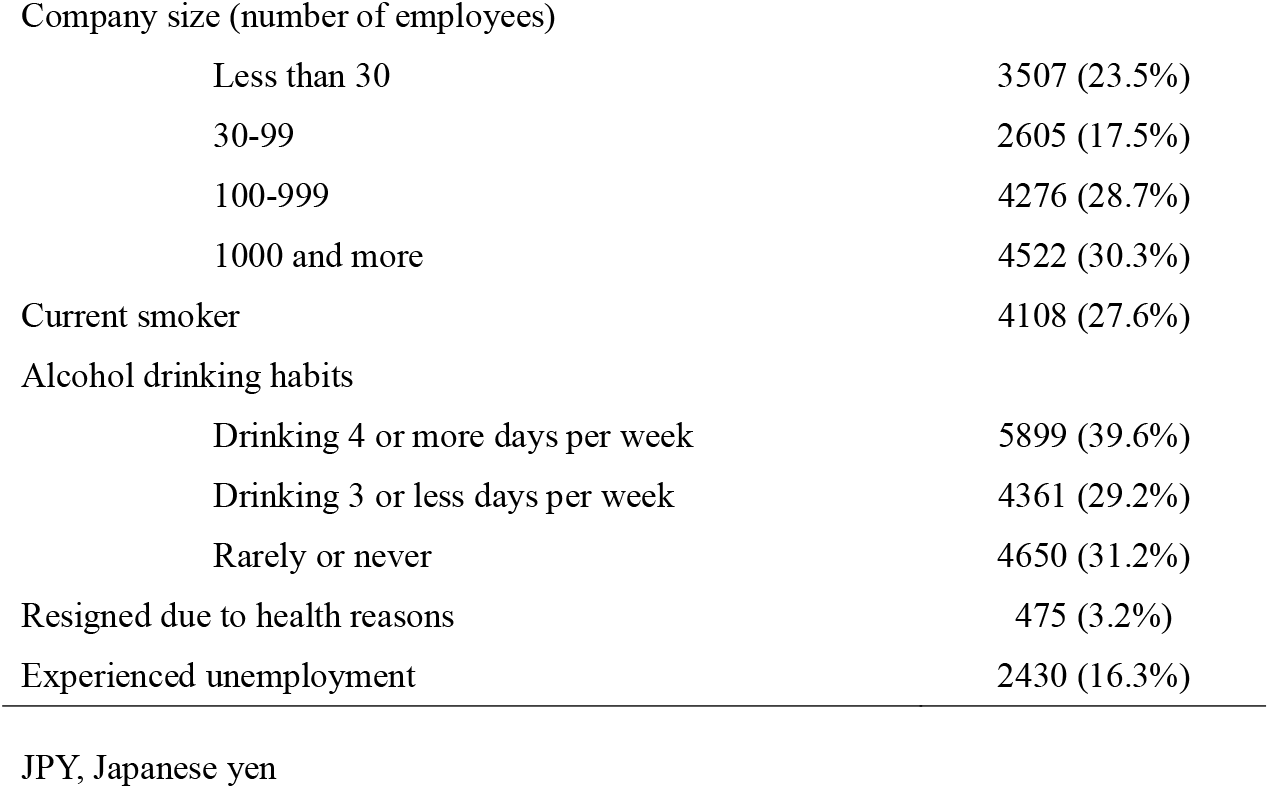
Baseline characteristics

|  | n (%) |
| --- | --- |
| Total | 14910 |
| Age, mean (SD) | 48.2 (9.9) |
| Sex, men | 8682 (58.2%) |
| Marital status |  |
| Married | 8638 (57.9%) |
| Divorced | 1449 (9.7%) |
| Never married | 4823 (32.3%) |
| Income (million JPY) |  |
| Less than 4.00 | 3574 (24.0%) |
| 4.00-5.99 | 3581 (24.0%) |
| 6.00-7.99 | 3089 (20.7%) |
| 8.00 and more | 4666 (31.3%) |
| Education |  |
| Junior high school | 178 (1.2%) |
| High school | 4047 (27.1%) |
| Vocational school/college, university, graduate school | 10685 (71.7%) |
| Job type |  |
| Desk work | 8411 (56.4%) |
| Work involving communicating with people | 3231 (21.7%) |
| Manual work | 3268 (21.9%) |

|  |  |  |
| --- | --- | --- |
| Company size (number of employees) |  |  |
| Less than 30 |  | 3507 (23.5%) |
| 30-99 |  | 2605 (17.5%) |
| 100-999 |  | 4276 (28.7%) |
| 1000 and more |  | 4522 (30.3%) |
| Current smoker |  | 4108 (27.6%) |
| Alcohol drinking habits |  |  |
| Drinking 4 or more days per week |  | 5899 (39.6%) |
| Drinking 3 or less days per week |  | 4361 (29.2%) |
| Rarely or never |  | 4650 (31.2%) |
| Resigned due to health reasons |  | 475 (3.2%) |
| Experienced unemployment |  | 2430 (16.3%) |

JPY, Japanese yen

Table 2 shows the associations between various symptoms and resignation or unemployment. For those participants who resigned for health reasons, the multivariate analysis showed that the OR of unemployment associated with pain-related problems was 2.21 (95% confidence interval (CI): 1.57–3.10), that associated with limited physical movement and mobility was 4.00 (95% CI: 2.73–5.86), that associated with fatigue, loss of strength or appetite, fever, dizziness, and moodiness was 3.08 (95% CI: 2.19–4.31), that associated with mental health problems was 2.94 (95% CI: 2.22–3.90), and that associated with sleep-related problems was 2.09 (95% CI: 1.47–2.98).

**Table 2.**
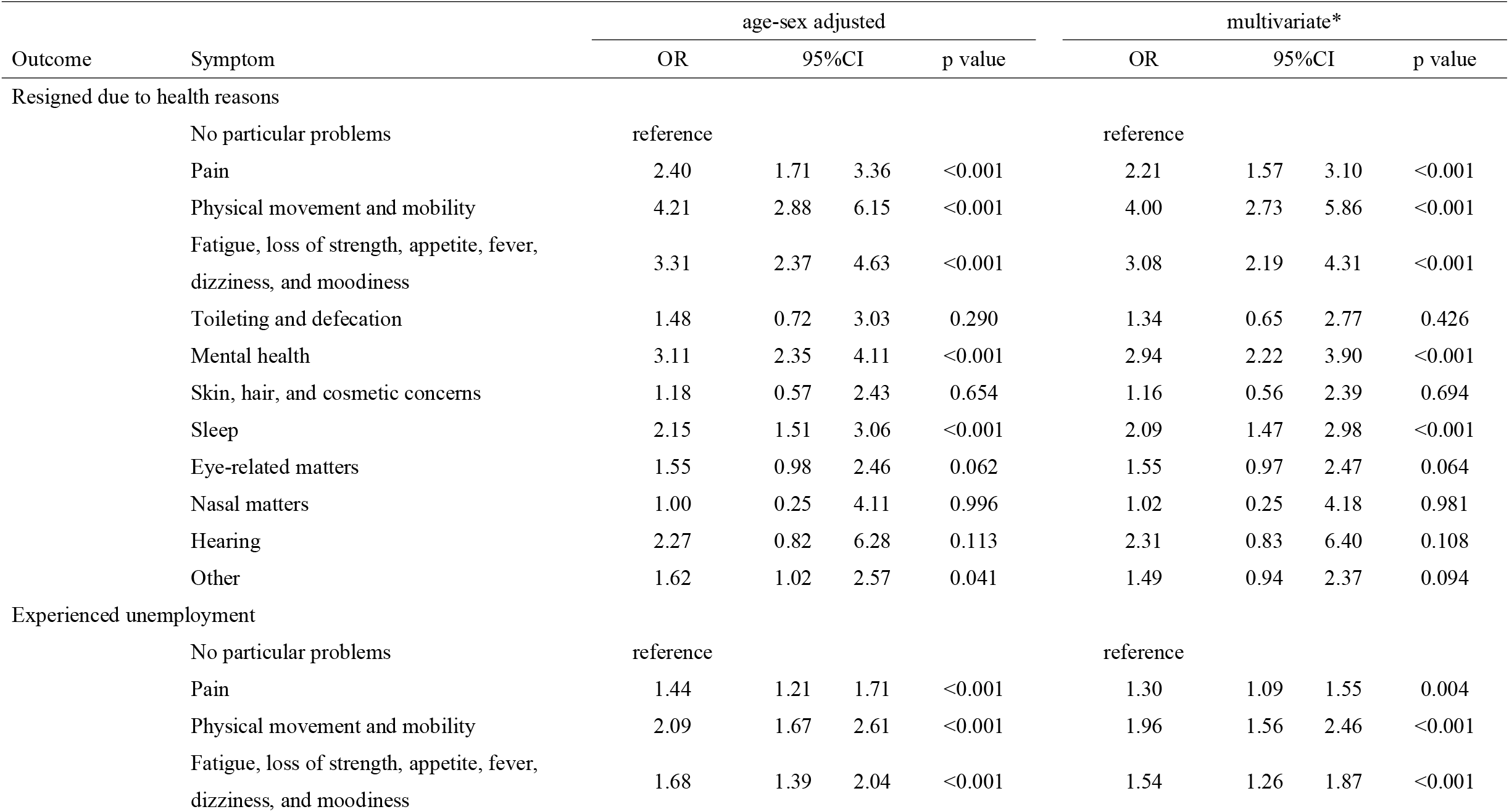
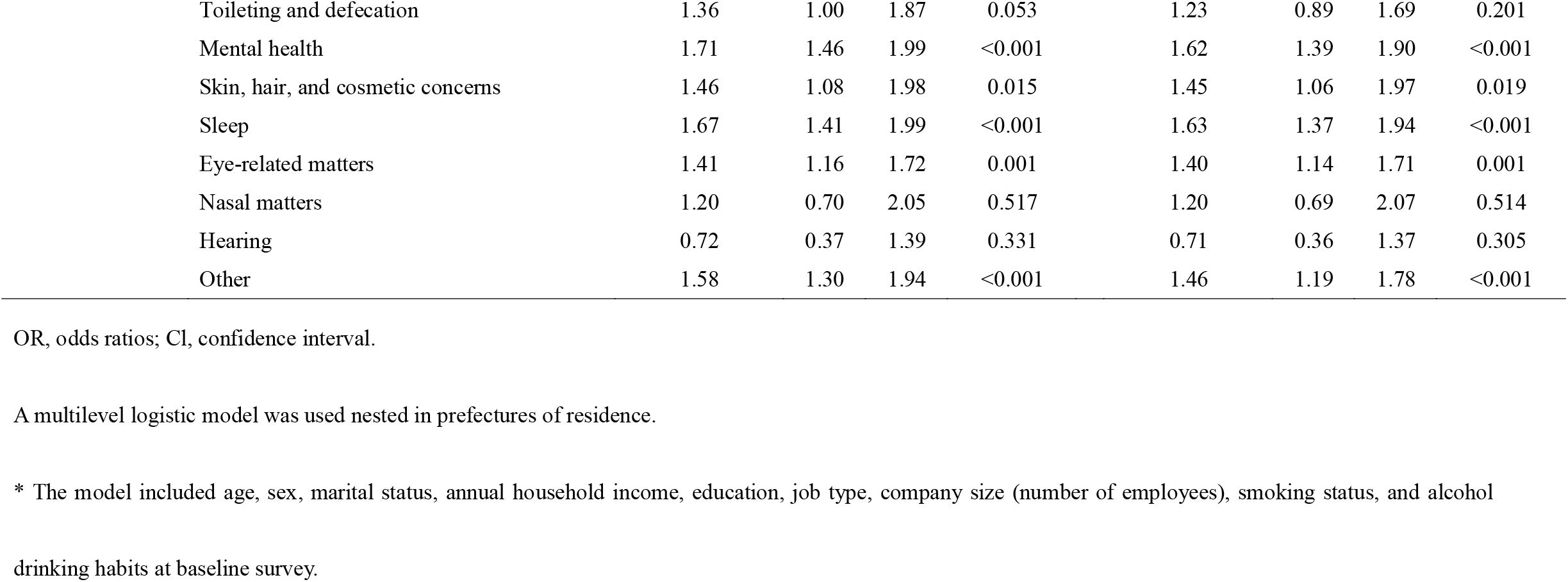
The associations between various symptoms and resignation or unemployment

Regarding unemployment, the multivariate analysis showed that the OR of unemployment associated with pain-related problems was 1.30 (95% CI: 1.09–1.55), that associated with limited physical movement and mobility was 1.96 (95% CI: 1.56–2.46), that associated with fatigue, loss of strength or appetite, fever, dizziness, and moodiness was 1.54 (95% CI: 1.26–1.87), that associated with mental health problems was 1.62 (95% CI: 1.39–1.90), that associated with skin, hair, and cosmetic concerns was 1.45 (95% CI: 1.06–1.97), that associated with sleep-related problems was 1.63 (95% CI: 1.37–1.94), and that associated with eye-related matters was 1.40 (95% CI: 1.14–1.71).

## Discussion

The results of this study showed that workers who had a specific symptom were at increased risk of either resigning or experiencing unemployment during the following year. The symptoms presenting the greatest risk to employees in terms of resigning or experiencing unemployment were mobility-related issues, followed by chronic fatigue and mental health problems.

There are several possible mechanisms by which workers with health-related symptoms either resign or experience unemployment. First, workers with health-related symptoms experience impaired work functioning^26^, resulting in inability to perform certain tasks, such as business trips, shift work, and heavy lifting. Workers who experience impaired work functioning often either reduce their working hours, change their job descriptions, or change their workplace. This can lead to employment-related disadvantages, long-term absence, and unemployment^27-32^. Second, workers with health problems have to reduce their working hours because of the need to attend medical appointments. Casual workers who do not have access to sick leave might have to leave their jobs to seek medical treatment. Third, workers with health problems can experience stigma in relation to their illness, which can lead to employment-related disadvantages and unemployment^33,34^. In particular, workers with infectious diseases, mental illness, or cancer are likely to experience discrimination, harassment, changes in job descriptions and workplaces, limited career advancement, and unemployment.

We examined the association between each symptom and resignation or unemployment in terms of the International Classification of Functioning, Disability and Health (ICF).

Workers with mobility-related issues experience limitations in relation to activities such as commuting or moving around the workplace, and also experience limitations in relation to the activities required in their job, such as carrying heavy objects and working at heights. Thus, workers with musculoskeletal disorders, which are one of the major causes of mobility-related issues, are more likely to experience unemployment^35,36^.

Fatigue, loss of strength or appetite, fever, dizziness, and moodiness are not related to a single disease or illness, but can present as symptoms of various diseases or illnesses. For example, they can occur in relation to infections, malignant neoplasms, or neuropsychiatric disorders. Workers with these symptoms experience limitations in relation to their work performance, regardless of their job description. In addition, it is often difficult for fellow workers to recognize these symptoms, making it difficult for sufferers to obtain support from their supervisors and colleagues. This lack of support, which is listed as an environmental factor in the ICF, can result in them either resigning or losing their job.

Pain-related problems include various types of pain such as local pain, systemic pain, acute pain, and chronic pain, and are a major cause of presenteeism^5-6^. It has been estimated that 80% of Japanese workers are living with some form of pain^37^, which is directly related to various occupational limitations. For example, workers with back pain experience significant limitations at work, especially in relation to commuting and physical movement. Workers who experience headaches are not only limited in their physical activity, but also experience limitations in relation to work requiring mental functions such as memory, attention, cognition, and emotion. It is also known that chronic pain increases patient’s anxiety and fear of pain, making them feel their pain more acutely, and reducing any pain-inducing physical activity. In addition, it is difficult for fellow workers to recognize that a worker is experiencing pain, leading to a lack of support from supervisors and colleagues.

Mental health problems include various mental impairments. Mental health problems impact workers’ activities that require mental functions such as memory, attention, cognition, and emotions. For example, Johnston et al. (2019) identified work disabilities caused by depression, including reduced ability to plan, make decisions, and execute tasks^38^. Workers with mental health problems have difficulty in obtaining support from supervisors and colleagues, and are likely to experience stigma and various disadvantages in the workplace.

Sleep-related problems result in fatigue and reduced strength, as well as impairment of mental functions such as memory, attention, and cognition^39^. It has been reported that workers with insomnia are at high risk of developing impaired work functioning^40^. In addition, workers with sleep-related problems might not receive understanding and consideration from their fellow workers because sleep-related problems are difficult to recognize.

The results of our study showed that eye- and hearing-related problems did not increase the risk of resignation and unemployment because visual and auditory impairments are likely to be easily recognized and accommodated by supervisors and colleagues. In addition, nasal problems did not increase the risk of resignation or unemployment, possibly because nasal functions are not required in many occupations. Toileting and defecation-related problems did not increase the risk of resignation or unemployment, possibly because although these functions are necessary for life support, they are not directly related to work performance in most cases.

The COVID-19 pandemic might have increased the risk of unemployment for workers with health problems for the following reasons. First, during the pandemic, numerous companies introduced measures preventing workers with symptoms from coming to work in an attempt to limit the spread of the virus. Therefore, it is possible that some workers did not reveal their symptoms, knowing that they would be shunned by colleagues. Second, people with underlying conditions were at higher risk of severe illness or death from COVID-19. Thus, workers with symptoms might have avoided work involving contact with other people for fear of infecting vulnerable colleagues, thereby increasing the risk of resignation or unemployment. Third, the deterioration in employment conditions as a result of the COVID-19 pandemic might have increased the risk of retirement or unemployment for workers with specific symptoms. Fourth, the COVID-19 pandemic has had a negative impact on people’s ability to continue disease management, exacerbating workers’ symptoms and increasing the risk of resignation or unemployment as a result of worsening conditions that impede work participation.

This study has some limitations. First, unemployment information was based on self-reporting, although we expected few memory errors and little recall bias regarding changes in employment over the relatively short timeframe (one year). Second, no information was available regarding the onset of symptoms, severity, or treatment status. Despite having the same symptoms, the impact on people’s risk of unemployment can differ depending on the severity of their illness and their treatment status. Third, this study did not include detailed job descriptions for the respondents. A worker’s ability to perform his or her job is determined not only by the type and severity of symptoms, but also by the nature and demands of the job, along with the support of co-workers and company systems^41,42^. Fourth, because this study was conducted during the COVID-19 pandemic, the impact of various symptoms on unemployment and retirement might have differed from that during non-pandemic periods, and thus further evaluation should be conducted over a longer period.

In conclusion, the results of this study indicate that workers with specific symptoms were at greater risk of resigning or experiencing unemployment during the following year, and different symptoms had different ORs. Therefore, preventive interventions to resolve or improve workers’ symptoms could prevent their resigning or becoming unemployed.

## Data Availability

The data supporting the findings of this study are available from the corresponding author, Tomohisa Nagata, upon reasonable request.

## Acknowledgments

This study was supported and partly funded by the research grant from the University of Occupational and Environmental Health, Japan (no grant number); Japanese Ministry of Health, Labour and Welfare (H30-josei-ippan-002, H30-roudou-ippan-007, 19JA1004, 20JA1006, 210301-1, and 20HB1004); Anshin Zaidan (no grant number), the Collabo-Health Study Group (no grant number), and Hitachi Systems, Ltd. (no grant number) and scholarship donations from Chugai Pharmaceutical Co., Ltd. (no grant number). The funder was not involved in the study design, collection, analysis, interpretation of data, the writing of this article or the decision to submit it for publication.

The current members of the CORoNaWork Project, in alphabetical order, are as follows: Dr. Akira Ogami, Dr. Ayako Hino, Dr. Hajime Ando, Dr. Hisashi Eguchi, Dr. Keiji Muramatsu, Dr. Koji Mori, Dr. Kosuke Mafune, Dr. Makoto Okawara, Dr. Mami Kuwamura, Dr. Mayumi Tsuji, Dr. Ryutaro Matsugaki, Dr. Seiichiro Tateishi, Dr. Shinya Matsuda, Dr. Tomohiro Ishimaru, and Dr. Tomohisa Nagata, Dr. Yoshihisa Fujino (present chairperson of the study group), and Dr. Yu Igarashi. All members are affiliated with the University of Occupational and Environmental Health, Japan.

We thank Geoff Whyte, MBA, from Edanz (https://jp.edanz.com/ac) for editing a draft of this manuscript.

## Disclosure

**Ethical approval**: This study was approved by the ethics committee of the University of Occupational and Environmental Health, Japan (reference nos. R2-079 and R3-006).

**Informed consent**: Informed consent was obtained in the form of the website.

**Registry and the registration no. of the study/trial**: N/A

**Animal studies**: N/A

